# Functional Disability, Health Care Access, and Mental Health in Ukrainians Displaced by the 2022 Russian Invasion

**DOI:** 10.1101/2023.12.15.23300031

**Authors:** Tarandeep S. Kang, Michael G Head, Ken Brackstone, Kateryna Buchko, Robin Goodwin

## Abstract

Armed conflict has negative mental health impacts on internally displaced people and refugees, however, much less is known of its effects upon the mental health of displaced people and refugees with disabilities. We use European data (N=9,679), collected via an internet health needs survey across April-July 2022, to examine the mental health impacts of the 2022 Russian invasion of Ukraine on the mental health of displaced people with disabilities during the first months of the war. Regression models separately examined associations between functional impairment (vision, walking, existing mental health condition), access to healthcare, welfare payments, and anxiety and sleep quality, controlling for a number of sociodemographic variables including preferred language spoken at home, as an indicator of Russian or Ukrainian cultural identity. The presence of pre-existing mental health condition, mobility and vision impairment were each associated with higher levels of anxiety and poorer sleep quality. The ability to access health services and social security payments was also associated with better sleep and lower levels of anxiety. Humanitarian and local authorities must ensure Ukrainian refugees and IDPs are reviewed for their mental health needs, with particular attention to those with known disabilities.

## Introduction

Warfare and armed conflict pose significant risks to mental health. A meta-analysis found that 13% of individuals exposed to warfare or armed conflict were likely to have mild forms of depression, anxiety, or PTSD (Post Traumatic Stress Disorder), 4% moderate disorder, and 5% had severe disorders (Charlson et al., 2019). Displaced people are at particular risk of mental health disorders (anxiety, PTSD, and depression (Peconga and Høgh Thøgersen, 2020; Porter and Haslam, 2005).

War in Ukraine began in 2014, with an initial Russian invasion of the Donbas and Crimea regions. In 2022, this became a full-scale invasion of the entire country. This war sparked considerable migration: data from The Migration Observatory (Walsh and Sumption, 2022) show as of 22 July 2022 more than 10 million border crossings out of Ukraine, and more than 6 million Ukrainian refugees across Europe as a result of the 2022 invasion. Surveys of Ukrainians internally displaced in the 2014 conflict found 55% of respondents were at risk of somatic distress, with a high prevalence of PTSD (32%), depression (22%), and anxiety (17%) (Cheung et al., 2019; Roberts et al., 2019). Data collected a few weeks after the 2022 invasion found 53% of respondents met criteria for general psychological distress, 54% for anxiety, 47% for depression, and 12% for insomnia (Xu et al., 2023). Further work suggested PTSD risk among Ukrainians was elevated by displacement, either internally or as a refugee (Ben-Ezra et al., 2023). Perhaps unsurprisingly therefore the WHO forecasts that by 2025, half of all Ukrainians will have mental health problems (Interagency Coordination Council on Mental Health Protection and Office of the President of Ukraine, 2023). This mental health burden has become a national health priority, with the First Lady of Ukraine Olena Zelenska initiating a National Program of Mental Health and Psychosocial Support (Office of the President of Ukraine, 2023).

Substantial previous research has shown that people with disabilities are likely to be at additional risk of negative mental health consequences from a broad range of disaster types, including armed conflict and warfare (Stough, 2009; Stough and Kelman, 2018). Researchers have raised particular concerns about the welfare and human rights of individuals with disabilities in the Ukraine conflict (Patwary et al., 2023; Ruškus, 2022). Geographical analysis of Ukrainian Government data from 2017 indicates that during the conflict in the Donbas region, individuals with disabilities were less likely to leave their homes, and where they were able to leave, moved shorter distances than individuals without disabilities (Mykhnenko et al., 2022). During the first wave of displacement, 2014-2022, the state treated all IDPs in the same way, despite multiple experiences of IDPs and their specific needs. In data collected shortly after the 2022 wide-scale invasion, the extent of functional disability in the Ukrainian population was associated with greater post-traumatic stress (Kang et al., 2023).

In this study we focus on three disabilities: poor visual health, restricted mobility, and pre-existing mental illness (Shakespeare, 2017). We use Europe-wide survey data from a large number of Ukrainian refugees displaced by the 2022 Russian invasion. We examine the association between functional disability, symptoms of anxiety, and sleep quality in people who were internally displaced or refugees. To our knowledge, this association has never been previously studied in the context of a large-forced migration to multiple European countries. We suggest several hypotheses. First, we anticipate that greater functional impairment will be associated with higher levels of anxiety (Frank et al., 2019; Turner et al., 2006) and poorer reported sleep quality (An and Joo, 2016; Ramos et al., 2014; Seow et al., 2020). Conversely, we predicted that adequate access to health services (Satinsky et al., 2019) will be as warm sociated with lower levels of anxiety and better sleep quality. Given that access to resources can act as a buffer against poor mental health (Hobfoll, 2014, 2012, 1989), we hypothesise that access to welfare payments would be associated with lower levels of anxiety and improved sleep quality. In line with previous research, we hypothesise that younger individuals and females (Faravelli et al., 2013) will be a greater risk of anxiety and older individuals and females will be at greater risk of poor sleep quality (Madrid-Valero et al., 2017). We compare individuals remaining in Ukraine (IDPs) with those who moved elsewhere in Europe. We hypothesise that the those who have moved elsewhere could be more likely to report more positive mental health (Ben-Ezra et al., 2023).

## Methods

### Recruitment and Sample

The study is based on the online cross-sectional survey (“Health Needs of Ukrainian Refugees and Internally Displaced Persons”: (https://www.the-ciru.com/resin-ukraine) (Head et al., 2022; Perelli-Harris et al., 2023). The study was advertised on Facebook between April 11 and July 15, 2022. Participants completed an online “health needs” survey using Qualtrics XM. Dissemination was conducted primarily using Facebook Ads Manager, with an advert describing the survey appearing on individuals’ Facebook timelines along with the survey link. This allowed us to direct the survey toward Ukrainian speakers aged 18 and over living in Ukraine or in any European country apart from Russia. Data from Ads Manager shows that the advert was seen by an estimated 1 million people during this period with the average respondent seeing advert 3.92 times in their feed. The survey was also shared by 1400 people on Facebook, reaching an estimated further 64,000 people.

The survey was advertised and conducted in Ukrainian. Native speakers translated the questions from English to Ukrainian, and other native speakers then reviewed the questions to ensure context and meaning was not lost. Participants provided informed consent after reading the study information. Ethical approval was received from the University of Southampton Ethics Committee (Institutional Review Board ID: 71890) and the Ethics Committee of the Psychology Department, University of Warwick (Ref: PSY_PGR_22-23/34).

In total, 10,216 participants completed the survey. Thirty-seven participants were removed for being under 18 years of age, and 500 participants were removed for not answering fully the measures relevant to this study. Therefore, the sample in the present study consists of 9,679 participants. The mean age of respondents was 43.11 (*SD* = 10.83; *Age Range =* 18-93), including 3933 IDPs and 5754 refugees.

## Measures

### Outcome variables

#### Mental health

Anxiety was measured using the GAD-2 (Kroenke et al., 2007; Plummer et al., 2016; Spitzer et al., 2006). Participants rated how often over the past 2 weeks they “felt nervous, anxious, or on edge” and “not able to stop or control worrying”; 0 = *not at all*, 3 = *nearly every day*); summed to form a single index (α = .76).

#### Sleep quality

Respondents indicated perceived level of sleep quality over the past week with a single item (1 = *poor*; 5 = *excellent)* (Snyder et al., 2018), and the frequency of nightmares over the past two weeks (1 = *not at all*, 4 = *nearly every day*).

### Displacement

To categorise displacement, a full list of countries was presented and participants indicated the country in which they currently were living. We distinguished between those located outside of Ukraine (refugees) and those inside the country (internally displaced).

### Health and disability

We followed a broad and widely used definition of disability that is not restricted to physical or sensory impairments, but explicitly also includes mental health problems (Shakespeare, 2017). Health and disability-related variables were therefore assessed by three items: difficulty with walking 500m (binary response: yes/no), vision (Likert scale; 1 = *very poor*; 5 = excellent), and the presence of previous a previous mental health condition (binary response yes/no).

### Access to resources

We separately examined access to healthcare and access to welfare payments (both binary variables: access /no access).

### Demographic variables

Participants indicated their age gender, and language spoken at home (Ukrainian, Russian, other). They also reported their education (high [university degree or higher] and lower); and previous settlement type (rural, urban). Participants’ current residence was then collapsed these into two categories: still in Ukraine (IDP) or now living in another European country as a refugee. Relationship status was assessed as a binary variable (single/ not).

## Data analysis

We began by computing Welch’s independent samples t-tests (Delacre et al., 2017) to examine differences in anxiety and sleep quality by gender. Next, we performed a pair of linear regression analyses with anxiety and sleep quality measures as separate outcomes. Predictors were entered hierarchically. The first block consisted of demographics (country of residence [internally displaced, elsewhere in Europe], primary language, current location as urban or rural, age, sex, and relationship status [single or not]). In the second block, we added disability-related variables (difficulty with walking, vision health, and the presence of a previous mental health condition). In the final block, we added variables measuring access to welfare payments and access to healthcare services.

Finally, we conducted a pair of Bayesian linear regressions, one for each outcome, using JASP version 0.18.1 (JASP Team, 2023; Love et al., 2019; Wagenmakers et al., 2023) and the BAS package (Clyde et al., 2022). As the recommended default (Rouder and Morey, 2012; van den Bergh et al., 2021), we make use of the Jeffreys–Zellner–Siow (JZS) prior (Liang et al., 2008; Zellner, 1986; Zellner and Siow, 1980). The Bayes Factor provides a measure of the strength of evidence supporting the alternative hypothesis compared to the null (Jeffreys, 2003). In this analysis we compare all combinations of predictors against the null.

## Results

Overall respondents reported high levels of anxiety (M, 3.59 SD 1.77) and poor sleep quality (M, 2.72, SD, .85). The majority (68.5%) of participants exceeded the GAD-2 cut-off score of 3 (Plummer et al., 2016). 42.2 % of respondents reported having nightmares on several days over the past two weeks, 16.4% on more than half the days, and 11.5% reported nightmares nearly every day. Women reported higher levels of anxiety (M 3.68 SD 1.73) than men (M 2.86 SD 1.89) *t*(1230.42) = -13.11, *p* = < .001. Women also reported poorer sleep (M 2.71 SD .85) than men (M 2.85 SD .85) *t*(1288.51) = 5.07, *p* = < .001 while older age was correlated with greater anxiety (r= .021, p = .05) and poorer sleep quality (r = -.068, p = <.001). Descriptive statistics for the key variables are in Table 1 and a full correlation matrix is presented in Table 2.

**Table 1:**
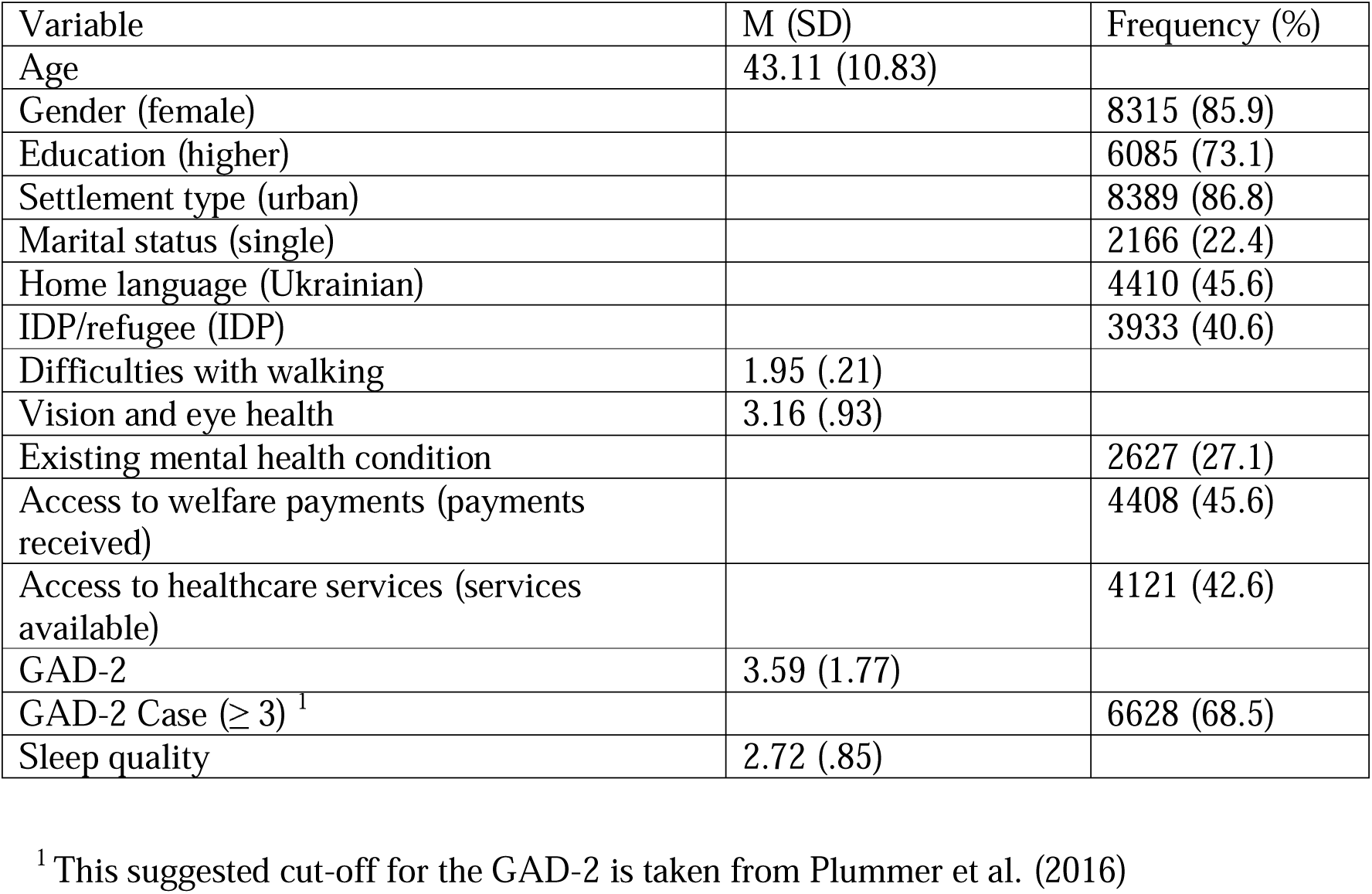
Descriptive statistics.

| Variable | M (SD) | Frequency (%) |
| --- | --- | --- |
| Age | 43.11 (10.83) |  |
| Gender (female) |  | 8315 (85.9) |
| Education (higher) |  | 6085 (73.1) |
| Settlement type (urban) |  | 8389 (86.8) |
| Marital status (single) |  | 2166 (22.4) |
| Home language (Ukrainian) |  | 4410 (45.6) |
| IDP/refugee (IDP) |  | 3933 (40.6) |
| Difficulties with walking | 1.95 (.21) |  |
| Vision and eye health | 3.16 (.93) |  |
| Existing mental health condition |  | 2627 (27.1) |
| Access to welfare payments (payments received) |  | 4408 (45.6) |
| Access to healthcare services (services available) |  | 4121 (42.6) |
| GAD-2 | 3.59 (1.77) |  |
| GAD-2 Case ($\geq 3$)<sup>1</sup> | | 6628 (68.5) |
| Sleep quality | 2.72 (.85) |  |

<sup>1</sup> This suggested cut-off for the GAD-2 is taken from Plummer et al. (2016)

**Table 2:** Correlation Matrix.

|  |  | Age | Gender | Home<br>Language | Urban/rural | Welfare<br>payments | Vision<br>and eye<br>health | Healthcare<br>Access | Difficult<br>walking<br>500m | Marital<br>status | Anxiety | Existing<br>Mental<br>Illness | Sleep<br>quality |
| --- | --- | --- | --- | --- | --- | --- | --- | --- | --- | --- | --- | --- | --- |
| Age | r | -- |  |  |  |  |  |  |  |  |  |  |  |
|  | p <. |  |  |  |  |  |  |  |  |  |  |  |  |
|  | N | 9174 |  |  |  |  |  |  |  |  |  |  |  |
| Gender | r | -.090** | -- |  |  |  |  |  |  |  |  |  |  |
|  | p <. | <.001 |  |  |  |  |  |  |  |  |  |  |  |
|  | N | 8854 |  |  |  |  |  |  |  |  |  |  |  |
| Language at<br>Home | r | .027* | -.031** |  |  |  |  |  |  |  |  |  |  |
|  | p <. | .011 | .003 |  |  |  |  |  |  |  |  |  |  |
|  | N | 8937 | 9100 |  |  |  |  |  |  |  |  |  |  |
| Urban/rural | r | -.029** | .025* | -.056** | -- |  |  |  |  |  |  |  |  |
|  | p <. | .006 | .015 | <.001 |  |  |  |  |  |  |  |  |  |
|  | N | 9083 | 9244 | 9329 |  |  |  |  |  |  |  |  |  |
| Welfare<br>Payments | r | .039** | .027* | .014 | -.019 | -- |  |  |  |  |  |  |  |
|  | p <. | <.001 | .010 | .174 | .058 |  |  |  |  |  |  |  |  |
|  | N | 9102 | 9260 | 9349 | 9502 |  |  |  |  |  |  |  |  |
| Vision and<br>eye health | r | -.274** | -.036** | -.002 | .036** | -.005 | -- |  |  |  |  |  |  |
|  | p <. | <.001 | <.001 | .819 | <.001 | .595 |  |  |  |  |  |  |  |
|  | N | 9164 | 9326 | 9411 | 9569 | 9585 |  |  |  |  |  |  |  |
| Healthcare<br>Access | r | .012 | -.052** | -.017 | -.007 | .098** | .070** | -- |  |  |  |  |  |
|  | p <. | .240 | <.001 | .102 | .526 | <.001 | <.001 |  |  |  |  |  |  |
|  | N | 9116 | 9278 | 9362 | 9518 | 9535 | 9600 | 9612 |  |  |  |  |  |
| Difficulty | r | -.132** | .075** | .024* | -.013 | .005 | .068** | .010 | -- |  |  |  |  |
| Walking | p <. | <.001 | <.001 | .019 | .218 | .625 | <.001 | .347 |  |  |  |  |  |
| 500m | N | 9171 | 9333 | 9419 | 9576 | 9593 | 9660 | 9609 | 9672 |  |  |  |  |
| Marital | r | .266** | .097** | -.014 | -.038** | .024* | -.091** | -.039** | -.040** | -- |  |  |  |
| Status | p <. | <.001 | <.001 | .178 | <.001 | .018 | <.001 | <.001 | <.001 |  |  |  |  |
|  | N | 9172 | 9335 | 9420 | 9577 | 9594 | 9661 | 9609 | 9669 | 9673 |  |  |  |
| Anxiety | r | .011 | .144** | -.006 | .016 | -.047** | -.137** | -.075** | -.031** | .002 | -- |  |  |
|  | p <. | .297 | <.001 | .555 | .118 | <.001 | <.001 | <.001 | .002 | .850 |  |  |  |
|  | N | 9113 | 9277 | 9356 | 9511 | 9526 | 9594 | 9543 | 9601 | 9602 | 9605 |  |  |
| Existing | r | .065** | .016 | .011 | -.003 | -.009 | -.144** | -.041** | -.004 | .041** | .233** |  |  |
| Mental | p <. | <.001 | .119 | .284 | .798 | .380 | <.001 | <.001 | .718 | <.001 | <.001 |  |  |
| Illness | N | 9174 | 9337 | 9423 | 9580 | 9597 | =9664 | 9612 | 9672 | 9673 | 9605 | 9676 |  |
| Sleep | r | -.061** | -.052** | .027* | -.012 | .052** | .168** | .071** | .062** | -.032** | -.377** | -.186** | -- |
| Quality | p <. | <.001 | <.001 | .010 | .243 | <.001 | <.001 | <.001 | <.001 | .001 | <.001 | <.001 |  |
|  | N | 9165 | 9330 | 9412 | 9569 | 9586 | 9653 | 9601 | 9661 | 9662 | 9595 | 9665 | 9665 |

\*\* . Correlation is significant at the 0.01 level (2-tailed).
\* . Correlation is significant at the 0.05 level (2-tailed).

Table 3 provides frequentist linear regression models for associations with anxiety and sleep quality. Female gender was significantly associated with anxiety, as was remaining in Ukraine. We also found that being single, having a pre-existing mental health condition, and greater difficulties with vision and mobility were also associated with greater anxiety. Finally, not having access to welfare payments or to healthcare was also associated with higher levels of anxiety. With regard to sleep quality, we found that those who are males, refugees, and who have higher levels of education report poorer sleep quality. We also find that speaking Russian at home is associated with poorer sleep. As with anxiety we again find that the presence of pre-existing mental health condition, together with greater difficulties with vision and mobility, and a lack of access to healthcare and welfare payments, are each associated with poorer sleep quality.

**Table 3.**
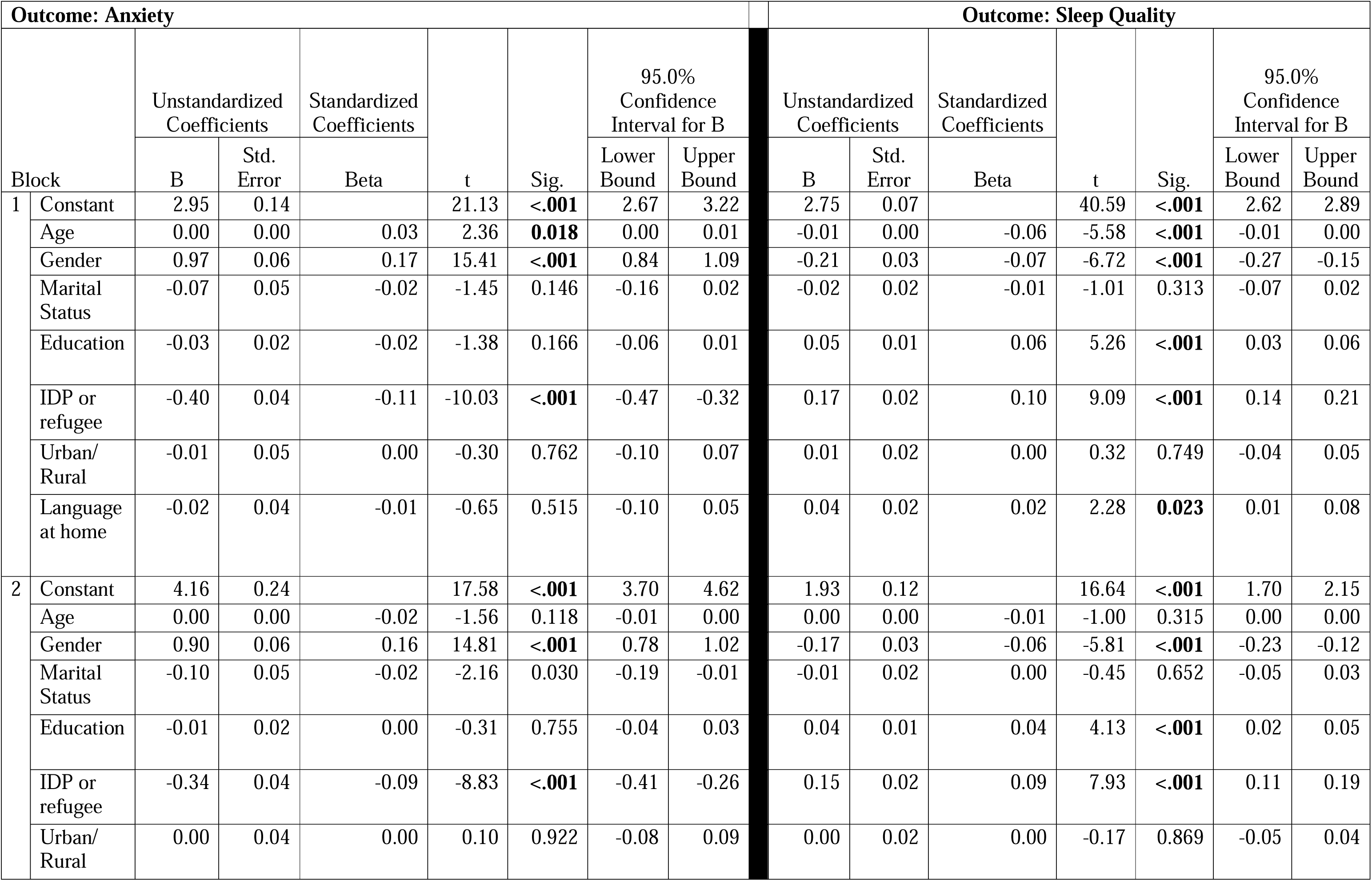
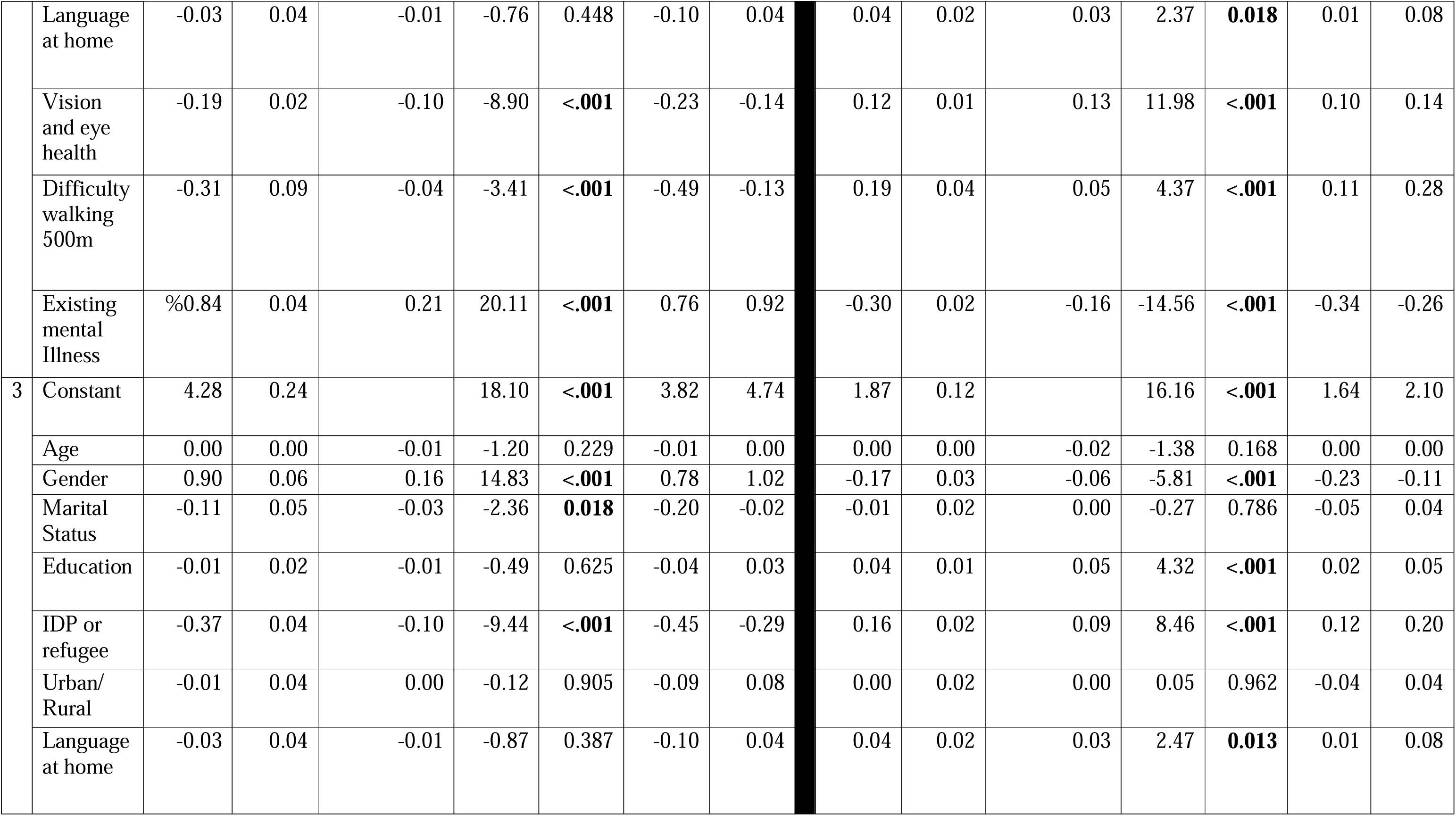
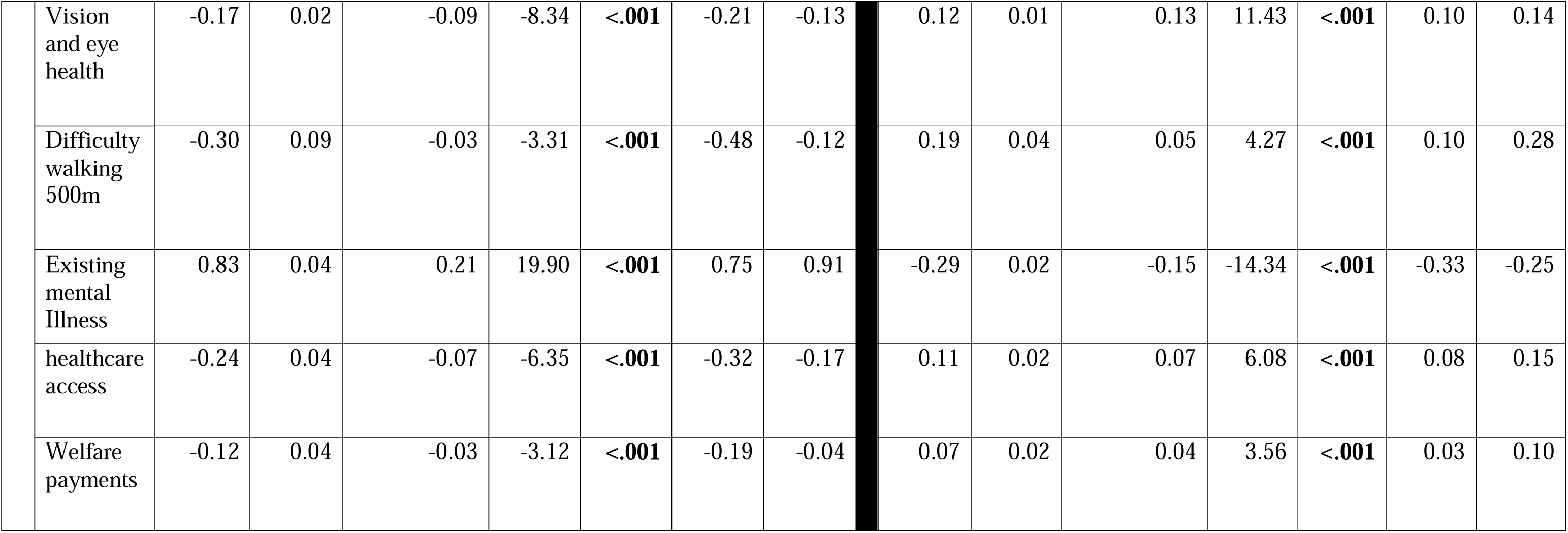
Linear Regression.

Turning to the results of our Bayesian regression models we find inclusion Bayes factors far in excess of 100 (Table 4 and 5) which in accordance with a widely used interpretation scheme (Jeffreys, 2003; Lee and Wagenmakers, 2014) means we have extremely strong evidence for the negative association between difficulties with walking, vision and pre-existing mental health conditions, with both anxiety and sleep quality. We also have extremely strong evidence that access to welfare payments and healthcare services is associated with lower levels of anxiety, higher levels of self-reported sleep quality.

**Table 4:** Posterior Summaries of Coefficients Predicting Anxiety.

| Coefficient | P(incl) | P(excl) | P(incl\|data) | P(excl\|data) | $BF_{inclusion}$ | Mean | SD | 95% Credible Interval | |
| --- | --- | --- | --- | --- | --- | --- | --- | --- | --- |
|  |  |  |  |  |  |  |  | Lower | Upper |
| Intercept | 1.000 | 0.000 | 1.000 | 0.000 | 1.000 | 3.591 | 0.018 | 3.557 | 3.628 |
| Age | 0.500 | 0.500 | 0.319 | 0.681 | 0.469 | $-8.237 \times 10^{-4}$ | 0.002 | -0.005 | $4.181 \times 10^{-4}$ |
| Gender | 0.500 | 0.500 | 1.000 | $-2.220 \times 10^{-16}$ | $5.903 \times 10^{+45}$ | 0.901 | 0.061 | 0.786 | 1.028 |
| Education | 0.500 | 0.500 | 0.187 | 0.813 | 0.230 | -0.002 | 0.009 | -0.028 | 0.007 |
| IDP or Refugee | 0.500 | 0.500 | 1.000 | $-2.220 \times 10^{-16}$ | $3.886 \times 10^{+18}$ | -0.369 | 0.039 | -0.440 | -0.286 |
| Urban/Rural | 0.500 | 0.500 | 0.167 | 0.833 | 0.200 | $-3.582 \times 10^{-4}$ | 0.018 | -0.043 | 0.036 |
| Marital Status | 0.500 | 0.500 | 0.801 | 0.199 | 4.025 | -0.094 | 0.062 | -0.188 | 0.000 |
| Language at Home | 0.500 | 0.500 | 0.223 | 0.777 | 0.287 | -0.007 | 0.022 | -0.072 | 0.016 |
| Difficulty Walking 500 m | 0.500 | 0.500 | 0.956 | 0.044 | 21.581 | -0.278 | 0.106 | -0.445 | 0.000 |
| Vision & Eye Health | 0.500 | 0.500 | 1.000 | $1.399 \times 10^{-14}$ | $6.976 \times 10^{+13}$ | -0.168 | 0.021 | -0.207 | -0.126 |
| Existing Mental Illness | 0.500 | 0.500 | 1.000 | $-2.220 \times 10^{-16}$ | $1.807 \times 10^{+82}$ | 0.824 | 0.042 | 0.746 | 0.909 |
| Healthcare Access | 0.500 | 0.500 | 1.000 | $1.727 \times 10^{-8}$ | $5.789 \times 10^{+7}$ | -0.241 | 0.038 | -0.312 | -0.164 |
| Welfare Payments | 0.500 | 0.500 | 0.948 | 0.052 | 18.348 | -0.111 | 0.045 | -0.181 | 0.000 |

**Table 5:**
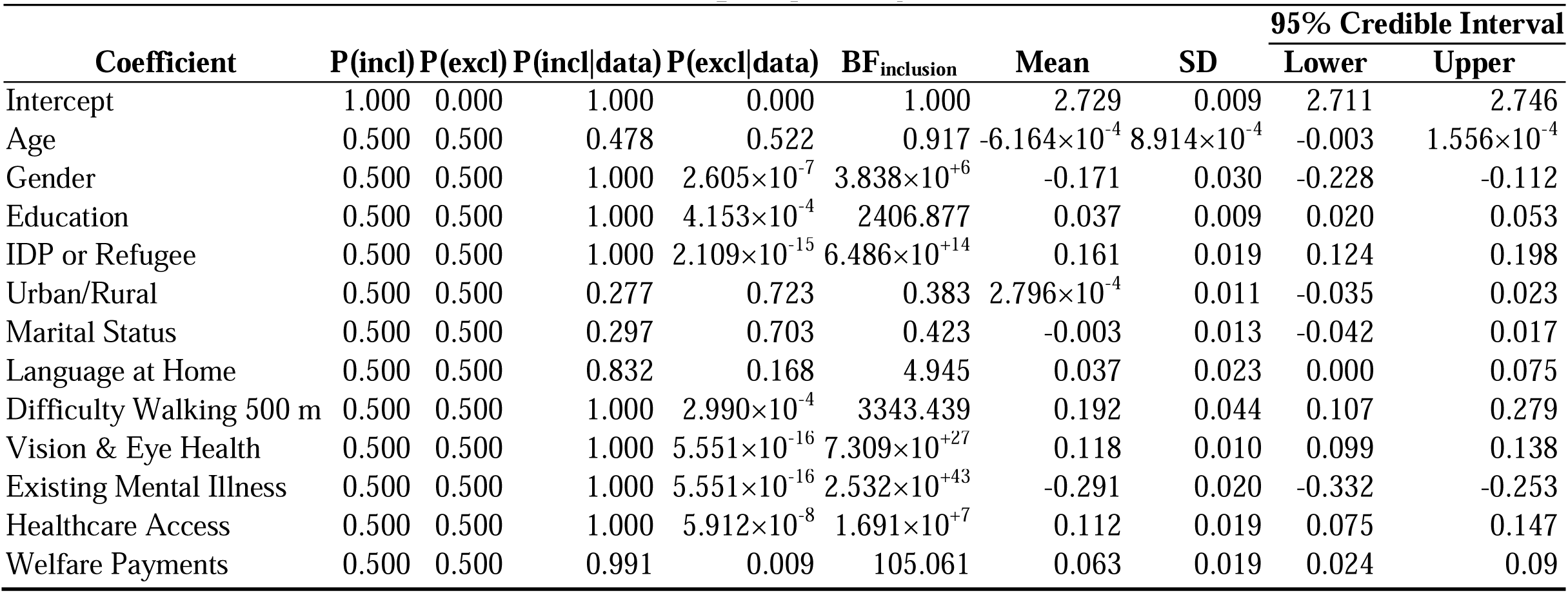
Posterior Summaries of Coefficients Predicting Sleep Quality.

| Coefficient | P(incl) | P(excl) | P(incl\|data) | P(excl\|data) | BF<sub>inclusion</sub> | Mean | SD | 95% Credible Interval |  |
| --- | --- | --- | --- | --- | --- | --- | --- | --- | --- |
|  |  |  |  |  |  |  |  | Lower | Upper |
| Intercept | 1.000 | 0.000 | 1.000 | 0.000 | 1.000 | 2.729 | 0.009 | 2.711 | 2.746 |
| Age | 0.500 | 0.500 | 0.478 | 0.522 | 0.917 | $-6.164 \times 10^{-4}$ | $8.914 \times 10^{-4}$ | -0.003 | $1.556 \times 10^{-4}$ |
| Gender | 0.500 | 0.500 | 1.000 | $2.605 \times 10^{-7}$ | $3.838 \times 10^{+6}$ | -0.171 | 0.030 | -0.228 | -0.112 |
| Education | 0.500 | 0.500 | 1.000 | $4.153 \times 10^{-4}$ | 2406.877 | 0.037 | 0.009 | 0.020 | 0.053 |
| IDP or Refugee | 0.500 | 0.500 | 1.000 | $2.109 \times 10^{-15}$ | $6.486 \times 10^{+14}$ | 0.161 | 0.019 | 0.124 | 0.198 |
| Urban/Rural | 0.500 | 0.500 | 0.277 | 0.723 | 0.383 | $2.796 \times 10^{-4}$ | 0.011 | -0.035 | 0.023 |
| Marital Status | 0.500 | 0.500 | 0.297 | 0.703 | 0.423 | -0.003 | 0.013 | -0.042 | 0.017 |
| Language at Home | 0.500 | 0.500 | 0.832 | 0.168 | 4.945 | 0.037 | 0.023 | 0.000 | 0.075 |
| Difficulty Walking 500 m | 0.500 | 0.500 | 1.000 | $2.990 \times 10^{-4}$ | 3343.439 | 0.192 | 0.044 | 0.107 | 0.279 |
| Vision & Eye Health | 0.500 | 0.500 | 1.000 | $5.551 \times 10^{-16}$ | $7.309 \times 10^{+27}$ | 0.118 | 0.010 | 0.099 | 0.138 |
| Existing Mental Illness | 0.500 | 0.500 | 1.000 | $5.551 \times 10^{-16}$ | $2.532 \times 10^{+43}$ | -0.291 | 0.020 | -0.332 | -0.253 |
| Healthcare Access | 0.500 | 0.500 | 1.000 | $5.912 \times 10^{-8}$ | $1.691 \times 10^{+7}$ | 0.112 | 0.019 | 0.075 | 0.147 |
| Welfare Payments | 0.500 | 0.500 | 0.991 | 0.009 | 105.061 | 0.063 | 0.019 | 0.024 | 0.09 |

## Discussion

The situation for people with disabilities in Ukraine was difficult even prior to the invasion, with individuals facing stigma and discrimination, a lack of social and economic participation, inadequate access to healthcare, rehabilitation, and welfare, amongst other challenges (Byndyu, 2017; Gazizullin and Solodova, 2019; Lipsmeyer, 2003). In our study of almost 10,000 displaced Ukrainians nearly 70% indicated a likely diagnosis of generalised anxiety disorder. Anxiety and poor sleep quality were positively associated with all 3 forms of disability assessed (visual, mobility and prior existing mental health problems). Access to healthcare facilities and welfare payments were positively associated with lower levels of anxiety and with better sleep quality. Female respondents were more vulnerable to symptoms of anxiety and poor sleep, as were IDPs compared with refugees.

The high prevalence of anxiety found in our study was consistent with other findings amongst Ukrainian refugees. A study of Ukrainian refugees in Germany (Buchcik et al., 2023) also found high prevalence of psychological distress, with this particularly high amongst females. This is also in line with much previous data on the mental health impacts of exposure to war and forced displacement (Blackmore et al., 2020; Charlson et al., 2019). We also found that pre-existing mental health conditions were associated with higher levels of anxiety and poorer sleep quality. This is also in line with previous research suggesting that pre-existing mental health is generally associated with poorer mental health in individuals exposed to armed conflict (Rozanov et al., 2019).

We find evidence that good healthcare access is associated with lower levels of anxiety. Again, this is in accordance with previous research that healthcare access (Lebano et al., 2020) and, indeed concerns about access to healthcare, may themselves be a source of anxiety for refugee populations (Strong et al., 2015). An interview-based study of Ukrainians forced to flee to the UK (Galpin et al., 2023) also suggests that access to healthcare is itself a particular cause of concern among refugees, and this may be exacerbated by a lack of cultural competence and of familiarity with the system. These factors have also been highlighted by other studies in the UK, Romania, and Norway amongst others (Labberton et al., 2023; Poppleton et al., 2023, 2022; World Health Organization Regional Office for Europe, 2023). In previous research with refugees and asylum seekers given the right to remain in the UK (Rowley et al., 2020), security of access to the benefits system helps to mitigate mental health problems. In line with this study and with resource-based accounts of stress and coping, we hypothesised that access to social welfare systems as a means to financial resources would help to mitigate symptoms of psychological distress. Our results support this hypothesis and we find evidence that access to social welfare benefits is associated with decreases in symptoms of anxiety and poor sleep quality However, it should be noted that negative attitudes towards receiving welfare payments appear prevalent across post-communist Europe (Lipsmeyer, 2003; Sätre, 2014). This may be particularly acute for recent forced migrants from Ukraine, who, in our dataset, were typically highly educated, and may prefer self-sufficiency (Galpin et al., 2023). This “useful migrant” narrative may help in part to counter racist attitudes in receiving countries towards “Eastern Europeans” as “benefit tourists” (Burrell and Schweyher, 2019; Fox et al., 2015). Thus, it is important for future researchers to not only examine the availability of welfare payments, but also respondents’ willingness to access them, and to contrast this with the prevailing sentiment from the surrounding local communities and policies in receiving countries towards finding gainful employment.

We find that each of the three domains of functioning, mobility, pre-existing mental health condition, and vision health were associated with increased levels of anxiety and poorer sleep quality. This replicates findings with general populations of visually impaired individuals not affected by conflict showing visual impairment is associated with increasing levels of anxiety (Frank et al., 2019; Zhang et al., 2023) and poor sleep (Leger et al., 1996), as is mobility impairment (Peterson et al., 2021; Smith et al., 2019). Associations between pre-existing mental illness and anxiety disorders have been established both in the general population and trauma exposed groups (Kroenke et al., 2007; Neria et al., 2010). Finally, previous research has also highlighted the cooccurrence of pre-existing mental health conditions and poor sleep quality in both general and trauma exposed populations (Ohayon et al., 1998; Short et al., 2018). The present study also supports conclusions from our previous work with Ukrainians displaced by the war (Kang et al., 2023) and highlights the importance of studying multiple impairments. It is also however possible that individuals with especially severe impairments opted not to move, and so were not captured in the sample of this study. It may be especially beneficial for future researchers to focus on a wide range of types (and severity of impairments) because the role of disability in forced migration research remains understudied.

We find that individuals who were internally displaced reported the highest levels of distress. This may obviously be because they are in greater danger than those individuals who been able to leave the country. We note too that those more resourced may have been more easily able to leave: wealthier and better-connected individuals (Abramitzky et al., 2022; Munroe et al., 2023; UNHCR, 2022), and those with better health (Mykhnenko et al., 2022) are among those who are more likely to move a greater distance and leave the country if possible. This study focused only on adults. Many Ukrainian children with disabilities live in residential institutions, these institutions are places of captivity, arbitrariness and humiliation. (Onufryk, Ukrainian Network of Child’s Rights, Annual report, 2022). Given this, the continuing war, mental health impacts upon children with disabilities displaced in Ukraine are likely to be even more severe than the adults we have surveyed in the present work. Future work should focus in particular on children who are displaced and forced to become refugees.

Insomnia poses a substantial problem to refugee populations (Baskaran et al., 2023). One reason that individuals who remain in Ukraine may be specifically vulnerable to higher rates of poor sleep quality is because war-related remembering is associated with insomnia (Basishvili et al., 2012). For IDPs within Ukraine, they may not just be remembering, but are actively exposed to potential sources of danger and trauma. It is interesting to note that we find Russian speakers are a greater risk of poor sleep quality. This is in line with previous research finding lower levels of resilience in this population (Goodwin et al., 2023) and may also be related to the strengthening of the Ukrainian identity (and therefore of the Ukrainian language) that has taken place in recent years owing to hostilities since 2014 (Arel, 2018; Chayinska et al., 2022; Eras, 2023; Kurapov et al., 2022).

We recognise several limitations in our study. The main data source, Facebook, of this study is both a strength and limitation. By using Facebook, we accessed a large sample of individuals who may have been otherwise hard to reach, and our number of participants greatly exceeds most other similar studies. Facebook has previously been shown to be useful in accessing such hard-to-reach populations (Iannelli et al., 2020; Kayrouz et al., 2016). On the other hand, the survey was explicitly advertised as a “health needs survey,” and individuals already more concerned than the average population about their health needs may be overrepresented in the sample. Relatedly, this study assessed only a limited number of functional impairments, which meant that it missed the full diversity of impacts of forced migration on other areas of functioning. Participants required access to the internet and a safe space to be able to complete the survey. Thus, we cannot be sure of how representative the survey is of the IDP and refugee population. Several of the measures we used required participants to select yes/no binaries, which may have caused to miss a finer grained picture of these impacts. Finally, this study was conducted only in Ukrainian, which may have ignored some of the linguistic and cultural complexity of the country (Barrington and Herron, 2004). This is especially important given that recent research has shown that differences in cultural identification (as a speaker of either Ukrainian or Russian) differentially impacts perceptions of national resilience (Goodwin et al., 2023).

There are several practical implications of our study. Real-time data in humanitarian situations are difficult to gather, and our large sample size provides useful and practical insight. Countries receiving refugees should be especially cognisant to provide them with timely and appropriate healthcare and welfare, as problems with accessing such services can themselves be a substantial source of distress, alongside the need to treat sources of distress and trauma, themselves. Individuals with poor vision may be particularly vulnerable to post-migration distress, and so clinicians may wish to pre-emptively engage with members of this group. Finally, as noted above, individuals who are most severely impaired in domains other than vision (intellectual impairment or those requiring full-time use of a wheelchair) may have particularly acute difficulties with migration, and so may have been unable to leave. It is therefore likely to be important for local authority, national-level governments, and humanitarian agencies to provide the Ukrainian state with additional medical assistance to better provide for the needs of IDPs who remain in the country, and for the refugee populations.

At the time of writing, the Ukraine displacement crisis is arguably the highest-profile crisis of its kind globally. However, there must also be continuing conversations around the plight of refugees in other settings, for example in West Africa with Burkina Faso refugees in Ghana (Inusah et al., 2023) or in East Africa with Ethiopia taking many refugees from Somalia and South Sudan (UNHCR, 2023). There is urgent need to pool the lessons learned from each of these displacements, and to globally and collectively manage their socio-economic and health impacts.

## Data Availability

Data will be shared upon reasonable request to the authors

